# Brain morphology mediating the effect of genetic risk variants on Alzheimer’s disease

**DOI:** 10.1101/2024.01.12.24301205

**Authors:** Esmee M. Breddels, Yelyzaveta Snihirova, Ehsan Pishva, Sinan Gülöksüz, Gabriëlla A.M. Blokland, Jurjen Luykx, Ole A. Andreassen, David E.J. Linden, Dennis van der Meer, Alzheimer’s Disease Neuroimaging Initiative

## Abstract

**INTRODUCTION:** Late-onset Alzheimer’s disease (LOAD) has been associated with alterations in the morphology of multiple brain structures and it is likely that disease mechanisms differ between brain regions. Coupling genetic determinants of LOAD with measures of brain morphology could localize and identify primary causal neurobiological pathways.

**METHODS:** Mediation and Mendelian randomization (MR) analysis were performed using common genetic variation, T1 MRI and clinical data collected by UK Biobank and Alzheimer’s Disease Neuroimaging Initiative.

**RESULTS:** Thickness of the entorhinal cortex and the volumes of the hippocampus, amygdala, choroid plexus and inferior lateral ventricle mediated the effect of *APOE* ε4 on LOAD. MR showed that a thinner entorhinal cortex, a smaller hippocampus and amygdala, and a larger volume of the choroid plexus and inferior lateral ventricles, increased the risk of LOAD as well as vice versa.

**DISCUSSION:** Combining neuroimaging and genetic data can give insight into the causal neuropathological pathways of LOAD.

## 1. Background

Late-onset Alzheimer’s disease (LOAD) is a progressive, neurodegenerative disease with a complex polygenic architecture (1) and an estimated broad heritability of 60-80% based on twin studies (2). LOAD is characterized by memory impairment and disturbances in mood and behaviour. A diagnosis is primarily based on the presence of clinical symptoms (3). However, before the onset of symptoms, the presence of biomarkers such as neurodegeneration could potentially be used for an early diagnosis of LOAD, but is currently mainly used in research settings (4-6). LOAD cannot be cured and available medications only slow down progression and relieve symptoms at best (3, 7). The discovery of effective disease-modifying treatments is complicated by the late diagnosis, when neurons have already degenerated to the point where the process is irreversible (8-10). The use of biomarkers for an early diagnosis is difficult because pathological hallmarks such as amyloid-β and tau are not specific enough to distinguish LOAD from normal ageing (4, 5).

LOAD has been associated with alterations in the structure of numerous cortical and subcortical brain regions (11). The spreading of neurodegeneration follows a distinct pattern, and it is likely that different disease mechanisms are involved at different stages. Presumably, only a subset of brain regions is involved in the core pathophysiology, whereas the remainder is affected by and involved in secondary disease processes. The disease has been shown to start in the middle temporal lobe, in the entorhinal cortex and the hippocampus through atrophy patterns using immunostaining and magnetic resonance imaging (MRI) (12-14). There are many hypotheses about LOAD primary pathology, mostly regarding accumulation of amyloid-β and tau pathology (15). Brain volume loss occurring later in the disease, such as that of frontal areas, might be the result of disruption of major signalling pathways due to the degeneration of the middle temporal lobe (13, 16), but could also be induced by the same processes that induced neurodegeneration in the entorhinal cortex and hippocampus (15).

Genome-wide association studies (GWAS) have identified dozens of common genetic variants associated with LOAD (17-19). However, the effect sizes of these variants are small and, even combined, explained only a small fraction of the heritability of LOAD (17). The strongest effect is seen for *APOE*, explaining 27.3% of the heritability (20). The *APOE* gene encodes for apolipoprotein E (apoE), a protein involved in cholesterol metabolism and transport, neuronal growth and repair, and immunoregulation (20). There are four isoforms of *APOE* (ε1-ε4) based on two single nucleotide polymorphisms (SNPs) at positions rs429358 and rs7412 (21).

The presence of one or two *APOE* ε4 alleles, increased the risk of LOAD 3 and 12 times respectively (22). *APOE* ε4 status is thereby one of the most common risk factors of LOAD and is used for stratification of populations in the clinical and research setting (23). Other genetic variants related to LOAD were linked to the immune system and lipid metabolism (17).

Neuroimaging techniques, genetics, and especially the combination of the two have been underutilized as tools for predicting both the occurrence and progression of LOAD. Coupling genetic determinants of LOAD with measures of brain morphology could localize and identify primary causal neurobiological pathways. The aim of the present study was to find causal pathways from genetic risk variants via brain morphology to LOAD. To accomplish this, mediation analysis was performed. First tested the association between the genetic variants and LOAD. Next, we aimed to estimate whether genetic risk variants of LOAD altered the morphology of certain brain regions, and whether this change in morphology, induced by the genetic risk variant, is related to LOAD.

## 2. Methods

### 2.1 Study population

To determine which brain regions mediate the effect of LOAD genetic risk factors, we performed mediation analysis using data from the UK Biobank (UKB) population cohort. To validate our findings, all analyses were repeated on a second cohort, namely the case-control cohort of the Alzheimer’s Disease Neuroimaging Initiative (ADNI).

The UKB consortium has collected genetic, lifestyle and health information of approximately 500,000 individuals aged 40 to 69 years at time of recruitment (24). A subgroup of participants was reinvited to a follow-up visit which included brain MRI scanning (25). The dataset used in the current study included 46,852 participants of whom brain MRI scans were available (Supplement Figure 1). We selected individuals who had been genotyped (n = 45,549) and were of white British ethnicity (n = 39,565), as determined by self-report confirmed by genetic principal components (field 22006 of the UKB dataset). Only individuals with white British ethnicity were included to increase genetic homogeneity. We excluded participants without parental LOAD status or age or age at death (n=643). Participants with missing genotypes for specific SNPs of interest were excluded in a model-specific manner. The final sample size ranged between 37,728 and 38,922 per analysis (Supplement Table 2).

The ADNI cohort (http://adni.loni.usc.edu/) is a public-private partnership with the primary goal of ADNI has been to test whether serial MRI, positron emission tomography (PET), other biological markers, and clinical and neuropsychological assessment can be combined to measure the progression of mild cognitive impairment (MCI) and early Alzheimer’s disease. The ADNI cohort, containing data from ADNI1, ADNIGO and ADNI2, had a sample size of 1,740 participants. Exclusion for the current study was based on the absence of MRI data (n = 672) or genotyping (n = 279), American Indian or Alaskan Native, Asian or more than one race (n = 9), or a diagnosis of MCI (n = 304). The final sample contained 476 subjects, of which 223 were LOAD cases and 253 cognitively normal controls (Supplement Figure 1).

A description and the corresponding UKB and ADNI field code(s) of all the variables used for this study are presented in Supplementary Table 1. The data was provided by the respective consortia. The UKB consortium obtained ethical approval from the North West Multi-centre Research Ethics and ADNI obtained ethical approval from the Ethical Committees of each institution where the work was performed (26). No extra regulations were required for the current study.

### 2.2 Ascertainment of Alzheimer’s disease

To maximize statistical power, we created a LOAD proxy score for the UKB to maximize statistical power, in accordance with previous genetic studies (17, 18, 27). Parental LOAD status and age or age at death were used to create the LOAD proxy score for each participant (17, 27). The LOAD proxy score ranged from 0 to 2. The sample obtained from UKB contained one participant diagnosed with LOAD (ICD code G30 and/or F00), who was given a score of 2. For each parent with a diagnosis of LOAD, the proxy score increased by 1. If the parent was never diagnosed as LOAD, the contribution to the LOAD proxy score was based on the age (or age at death) of the parent ((100-age)/100). This contribution based on age was added to account for the possibility of the parent still being at risk of developing LOAD. Participants whose parent(s) were relatively young or died at a young age would obtain a too high LOAD proxy score. To limit the effect of young parents, the maximum contribution of a parent without LOAD was set to 0.32, the maximum population prevalence for AD, based on the prognosis of Hebert et al. (17, 28). Information on parental LOAD status and age (at death) was self-reported by the participants. For more information, see Jansen et al. (17). The AD proxy score was not normally distributed. A simulation with 100,000 replications concluded that the type one-error rate was preserved, and linear regression models were appropriate. The average (±SD) LOAD proxy score was 0.66 (±0.41).

The ADNI cohort contained memory-related diagnostic information, categorized as LOAD, MCI or cognitively normal, based on Mini-Mental State Examination (MMSE), Clinical Dementia Rating (CDR) and NINCDS/ADRDA Alzheimer’s criteria (29). The most recent diagnosis available per subject was used and only LOAD cases and cognitively normal controls were included.

### 2.3 Genotyping

Detailed information on the genotyping process of UKB (30) and ADNI (website: https://adni.loni.usc.edu/data-samples/data-types/genetic-data/) have been reported elsewhere. Briefly, 488,377 UKB participants were genotyped for 805,426 markers using the UK BiLEVE Axiom Array (49,950 participants) and the UKB Axiom Array (438,427 participants). We made use of the UKB v3 imputed data, based on the Haplotype Reference Consortium (HRC) reference panel. The BGEN file obtained from UKB was converted to PLINK binary format. SNPs with more than 10% missingness and SNPs failing the Hardy– Weinberg equilibrium test at p < 1 × 10−9 were filtered out.

Genotype data from the ADNI sample were obtained from the ADNI LONI database. Whole genome genotyping was performed using different arrays depending on the phase of recruitment. The Illumina Human610 Quad Beadchip and Illumina HumanOmniExpress Beadchip were used for ADNI1 and ADNIGO/2, respectively. The 1000 Genomes Phase 3 reference panel was used for imputation. Based on qualitative assessment of clustering of the first two principal components, non-European cases were removed. Quality control (QC) and imputation processes were performed according to published recommendations (31). Briefly, QC included removal of individuals with more than 10% missingness and variants deviating from Hardy-Weinberg equilibrium < 1e-3, or with low imputation quality (r2<.03).

### 2.4 Selection of genetic risk variants

Our initial analysis was performed on UKB, therefore UKB specific GWAS summary statistics were obtained from Jansen et al. (17) and used as input for the SNP2GENE function in the online platform Functional Mapping and Annotation of Genome-Wide Association Studies (FUMA), with default settings for the clumping was used (32). Based on the UKB release 2b 10,000 European reference panel population and UKB specific GWAS summary statistics, 13 genomic risk loci were identified and the lead SNP for each locus was selected (Supplement Table 2). Genotype information on these 13 lead SNPs of interest was extracted for all participants. No information could be obtained for rs115674611, rs184384746 and rs187370608 in either UKB or ADNI. Analyses were performed on the remaining 10 SNPs and *APOE* ε4 status. To calculate the number of *APOE* ε*4* alleles per participant, information on rs429358 and rs7412 was extracted for each participant. The nucleotide at rs429358 and rs7412 can be either cysteine (T) or arginine (C), and their combination leads to four *APOE* alleles: ε1, rs429358 (C) + rs7412 (T); ε2, rs429358 (T) + rs7412 (T); ε3, rs429358 (T) + rs7412 (C); and ε4, rs429358 (C) + rs7412 (C) as described previously (33)..

### 2.5 MRI data acquisition and pre-processing

T1-weighted MRI scans for UKB were acquired at three different sites, all using identical 3 Tesla Siemens Skyra scanners (Siemens Healthineers, Erlangen, Germany), with a 32-channel head coil and identical protocols (25). Images were sent to the imaging centre, where they were reconstructed and pre-processed using an automated pipeline. Pre-processing included corrections for head motion and other artefacts, and automated quality control for identifying equipment issues such as coil failure. Detailed information on the MRI acquisition and processing was published elsewhere (34).

ADNI T1-weighted MRI protocols can be found at http://adni.loni.usc.edu/methods/documents/mriprotocols/. In short, all scans were acquired using MRI scanners from GE Healthcare, Philips Medical Systems, or Siemens Medical Solutions. Images were sent to the Mayo Clinic for data correction and quality control procedures.

The current study included brain morphology measures, made available by UKB and ADNI, of surface area and mean thickness of 34 cortical regions, and volume of 25 subcortical, ventricular, callosal and other automatically segmented non-cortical regions. Cortical reconstruction and volumetric segmentation for all images (UKB and ADNI) was performed using FreeSurfer software (35), versions v5.3 (UKB) and v5.1 (ADNI1/2/GO). Measures from the left and right hemisphere were summed for measures of surface area and volume and averaged for thickness. In addition, intracranial volume (ICV), total surface area and global mean thickness were extracted. ICV was available for both UKB and ADNI, whereas total surface area and global mean thickness could only be extracted for UKB. For ADNI participants, total surface area was calculated as the sum of the surface areas of the 34 cortical regions included in the study. The global mean thickness was a weighted sum of the mean thickness of the 34 cortical regions, where the contribution of each region was determined by the proportion of the region’s surface area of the total surface area. All brain measures were mean-centred and standardized, i.e., converted to Z-scores, before inclusion in the models.

### 2.6 Statistical analysis

All analyses were performed in R version 4.1.2 (36). Data from ADNI were retrieved via the *ADNIMERGE* package (37) obtained from (https://ida.loni.usc.edu/login.jsp).

#### 2.6.1 Mediation analysis

The relationship between genetic variation, brain morphology and LOAD was assessed using mediation analysis (Figure 1) (38, 39) implemented in the *mediation* package (40) in R We constructed models whereby path “a” represents the relationship between the LOAD risk variant and brain morphology. Path “b” represents the relationship between brain morphology and LOAD, adjusted for genetic variation. Lastly, path “c” represents the total effect of the genetic variant on LOAD. The total effect is composed of the direct effect of the genetic variant on LOAD, adjusted for brain morphology (path “c”), and the indirect or mediation effect (path “ab”). The estimate of the mediation effect is the product of path “a” and path “b” and its significance was determined through bootstrapping. Age, sex, and site for UKB were included as covariates. We additionally included either total surface area, global mean thickness or total ICV as a global correction, for the analyses of the regional surface area, thickness or non-cortical volume, respectively. Due to the large number of assessment sites for ADNI and the small sample size per site, site could not be included as a covariate, in line with previous studies using this data. ADNI has standardised MRI protocols to minimize potential site and scanner effects, and no significant variability of MRI performance between sites has been previously shown (26).

**Figure 1:**
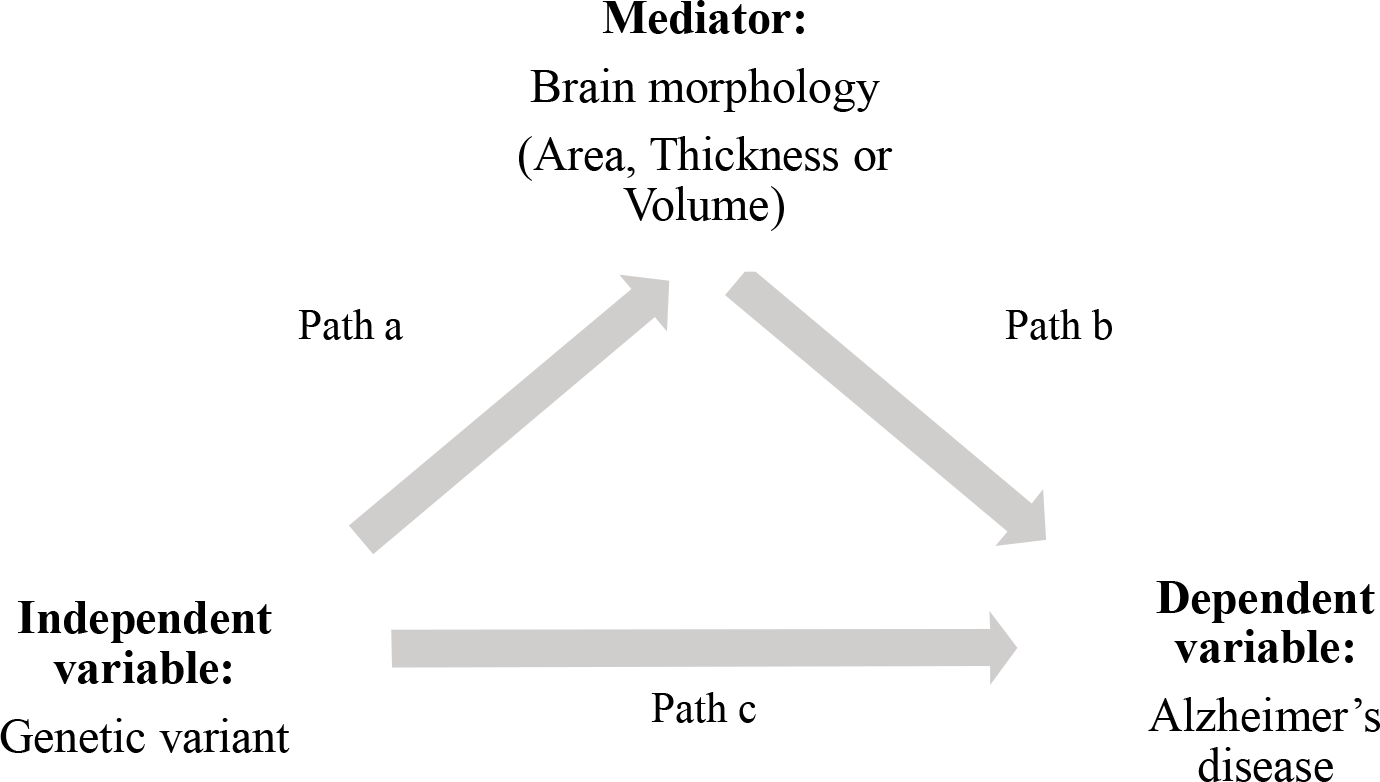
Mediation model. Path a represents the association between the genetic variant and measure of brain morphology (mediator model). Path b represents the association between brain morphology and LOAD. Combining path a and path b gives the mediation effect. Path c represents the total effect of the genetic variant on LOAD.

The analyses resulted in 1023 models for UKB and 1023 models for ADNI. Multiple comparisons correction was performed using the False Discovery Rate (FDR) set to 5%. Brain measures as well as the genetic variants were correlated, therefore the effective number of independent tests (41) was used in the FDR correction instead of the tests performed. The formula for FDR was adjusted by Li and Ji (41) to use the effective number of independent tests rather than the actual number of tests. The adjusted formula was used to calculate a significance threshold for each test.

Combining the results from our analysis on UKB and ADNI can potentially reveal new associations which might be borderline non-significant in the individual analysis. Therefore, the mediation effects obtained from the analysis on UKB and on ADNI were pooled using the inverse-variance weighted (IVW) method for meta-analysis (42). The standard errors of the mediation analysis estimates were approximated using the width of the 95% confidence intervals obtained via bootstrapping (5000 bootstrap simulations).

#### 2.6.2 Mendelian randomization

Mendelian randomization (MR) was used to determine the direction of causality, whether the change in brain morphology is cause or consequence of LOAD. MR is based on the assumption that if for example LOAD is caused by a change in a certain brain region, than any variable (e.g. genetic variants) influencing this region should also influence LOAD (43, 44). All 10 SNPs (see Supplementary Table 2) and *APOE* ε4 status were selected as instrumental variables. Only brain measures that were significant mediators were used as exposures.

MR was performed using the *MendelianRandomization* package (45). To minimize the risk of bias due to violation of assumptions, several MR methods were applied and compared for consistency of results. The MR methods used were the IVW method (46) (robust when all genetic variants are valid and horizontal pleiotropy is absent), weighted median (47) (robust to violations of assumptions by maximum half of the instrumental variables and outliers), and MR-Egger (48, 49) (robust to violation of the horizontal pleiotropy assumption by all instrumental variables). The validity of the results was additionally assessed by the intercept of the MR-Egger method (measure of pleiotropic effects (48)), I2 statistic (measure of instrument strength MR-Egger (47, 48)), Cochran’s Q statistic (measure of heterogeneity between variant-specific causal estimates (44)), and leave-one-out analysis (assessing the contribution of each genetic variant to the MR analysis (44)).

## 3. Results

### 3.1 Demographics

The UKB sample of 38,922 participants had a mean (± standard deviation; SD) age at the time of MRI of 64.4 (± 7.7) years and the distribution over males and females was 47.7% and 52.3% respectively (Table 1). In the ADNI dataset, the group of LOAD cases had more males (62.3%) compared to the control group (46.1%) (Table 1). The mean age (± SD) was 76.1 (± 7.74) years in the LOAD group and 76.2 (± 6.79) years for controls.

**Table 1:** Demographics of the UK Biobank and ADNI participants.

|  | UKB |  | ADNI |  | p-value |
| --- | --- | --- | --- | --- | --- |
|  |  | p-value | Cases | Controls |  |
| Age (years (SD)) | 64.4 (7.7) | 0.479 | 76.1 (7.74) | 76.2 (6.79) | 0.866 |
| Male (%) | 47.7 | <.0001 | 52.7 | 47.3 | 0.012 |
| Female (%) | 52.3 |  | 40.7 | 59.3 |  |

### 3.2 Associations between genetic variants and LOAD (path c)

*APOE* ε4, rs75627662 and rs7384878 were significantly associated with LOAD in both UKB and ADNI. Five additional SNPs were significantly related to LOAD in UKB participants. No additional SNPs were significantly associated with LOAD diagnosis in ADNI. The identified associations are summarized in Supplementary Table 3.

### 3.3 Associations between genetic variants and brain measures (path a)

Analysis on UKB data revealed 29 associations between genetic risk factors and brain regions (Supplementary Table 4). Most associations were found for *APOE* ε4, which was related to 9 brain measures. In the ADNI subset, significant associations of *APOE* ε*4* with the mean thickness of the entorhinal cortex, and the volumes of the amygdala, hippocampus and inferior lateral ventricle were found (Figure 2, Table 2). Additionally, there was a significant association between rs75627662 and entorhinal thickness, amygdala and hippocampal volume, and between rs7384878 and putamen volume.

**Figure 2:**
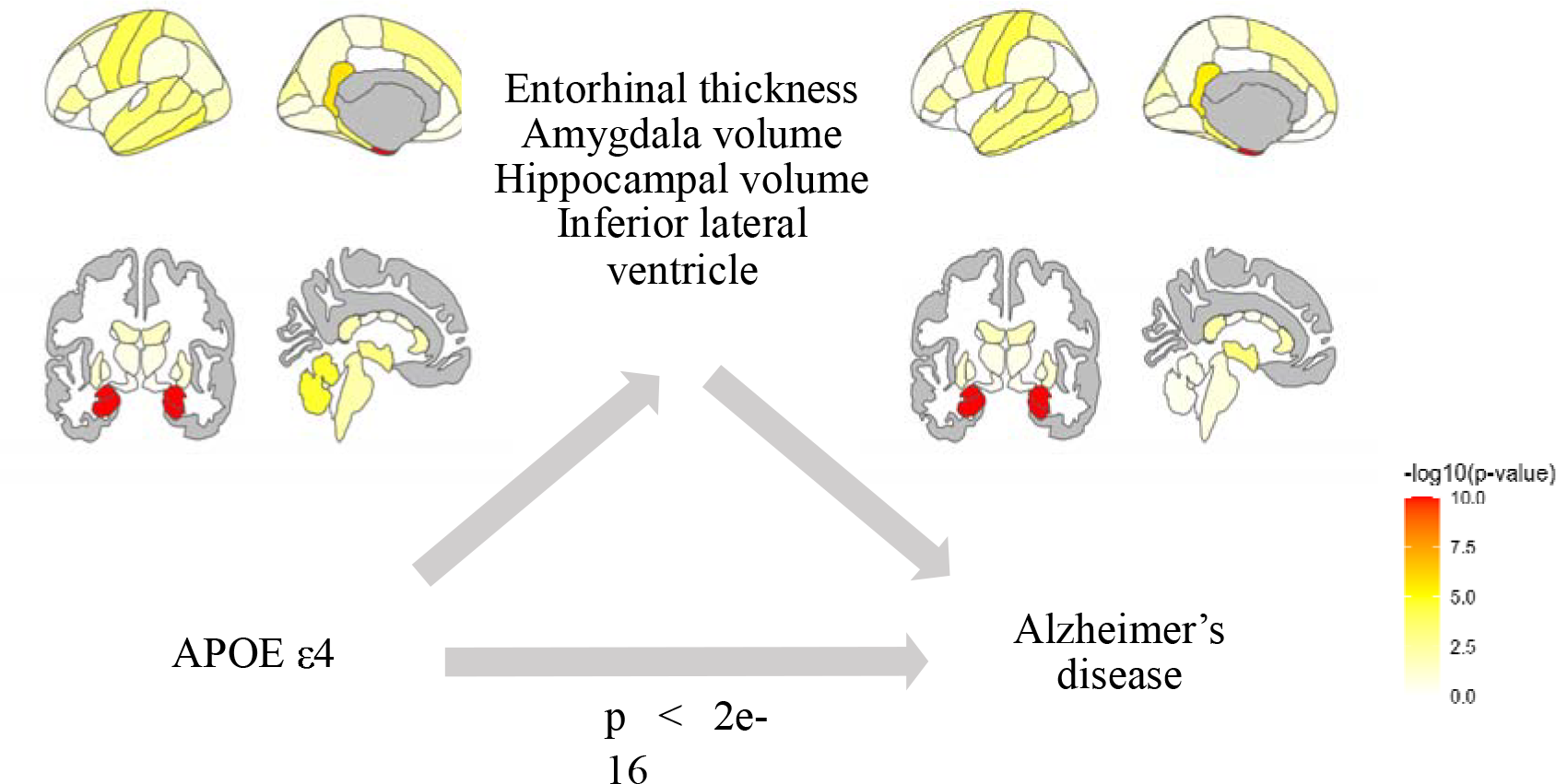
Results mediation analysis with APOE ε4. The effect of APOE ε4 was mediated by the entorhinal cortex, amygdala, hippocampus and inferior lateral ventricle.

**Table 2:**
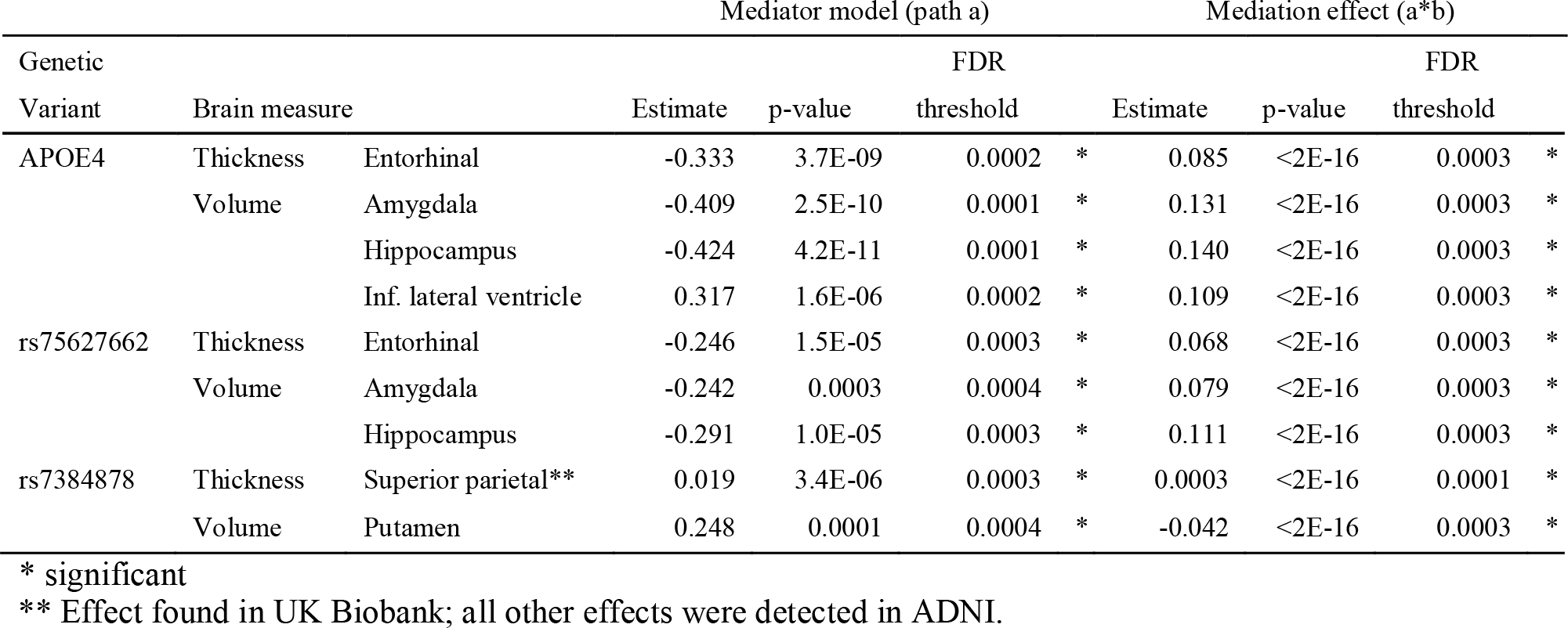
Significant results mediation analysis. The mediator model (path a) represents the effect of the genetic variant on the brain measure. The mediation effect (a*b) is the ACME of the brain measure on the association between the variant and LOAD. FDR is the p-value after FDR correction.

### 3.4 Mediation effect of brain measures (path a*b)

In the UKB dataset, we found a single mediation effect: the effect of rs7384878 on the LOAD proxy score was mediated by the thickness of the superior parietal cortex. A cytosine at rs7384878 compared to thymine was associated with a thicker superior parietal cortex, mediating the association between rs7384878 and an increased LOAD proxy score.

In ADNI, brain regions that were significantly related to genetic variants in path *a*, were also the mediators of the effect of the genetic variant on LOAD (Figure 2). Entorhinal thickness and the volumes of the amygdala, hippocampus, inferior lateral ventricle and choroid plexus significantly mediated the effect of *APOE* ε4 on LOAD (Table 2). Entorhinal thickness, hippocampal and amygdala volume additionally mediated the effect of rs75627662 on LOAD. The volume of the putamen mediated the effect of rs7384878 on LOAD. Looking at the directionality of the effects, *APOE* ε4 and rs75627662 were associated with thinner entorhinal cortex and smaller volumes of the amygdala and hippocampus, relating to a higher risk of LOAD. *APOE* ε4 allele count was associated with larger volume of the inferior lateral ventricles and choroid plexus, also associated with a higher risk of LOAD. Rs7384878 was associated with a higher volume of the putamen and decreasing risk of LOAD.

Combining the effects in the UKB and the ADNI by means of meta-analysis did not result in any significant effect (Supplementary Table 6). Closer inspection of the results showed that mediation effects that were significant in one cohort, were highly non-significant or in the opposite direction in the other cohort, see Supplementary Table 4.

### 3.5 Mendelian randomization

Mendelian randomization analysis was performed to assess the directionality of the associations between the mediating brain regions and LOAD. A significant bidirectional relationship was found between LOAD and the entorhinal cortex, amygdala, hippocampus, inferior lateral ventricle and choroid plexus (Supplementary Table 5; Figure 3). All MR methods (IVW, weighted median and MR-Egger) resulted in a significant negative effect of the entorhinal cortex, amygdala and hippocampus on LOAD. The reverse MR also resulted in a significant negative effect of LOAD on these brain regions. The relationship between the inferior lateral ventricle and choroid plexus with LOAD was significantly positive in both directions indicating that a larger volume of the inferior lateral ventricle and choroid plexus increased the risk of LOAD and vice versa. Sensitivity analysis revealed violations of assumptions in some of the models. The MR-Egger estimate was unreliable for the effect of the entorhinal cortex, choroid plexus, inferior lateral ventricle, hippocampus and amygdala (all I2 < 80%) on LOAD. However, for each analysis, the results across methods were in agreement on the directionality of the effects, supporting the conclusions. For the relation between the putamen and LOAD, only the IVW method was significant in both directions. The model testing the effect of putamen volume on LOAD gave a significant Cochran’s Q statistic (p <0.001) indicating a potential violation of the instrumental variable assumption.

**Figure 3:**
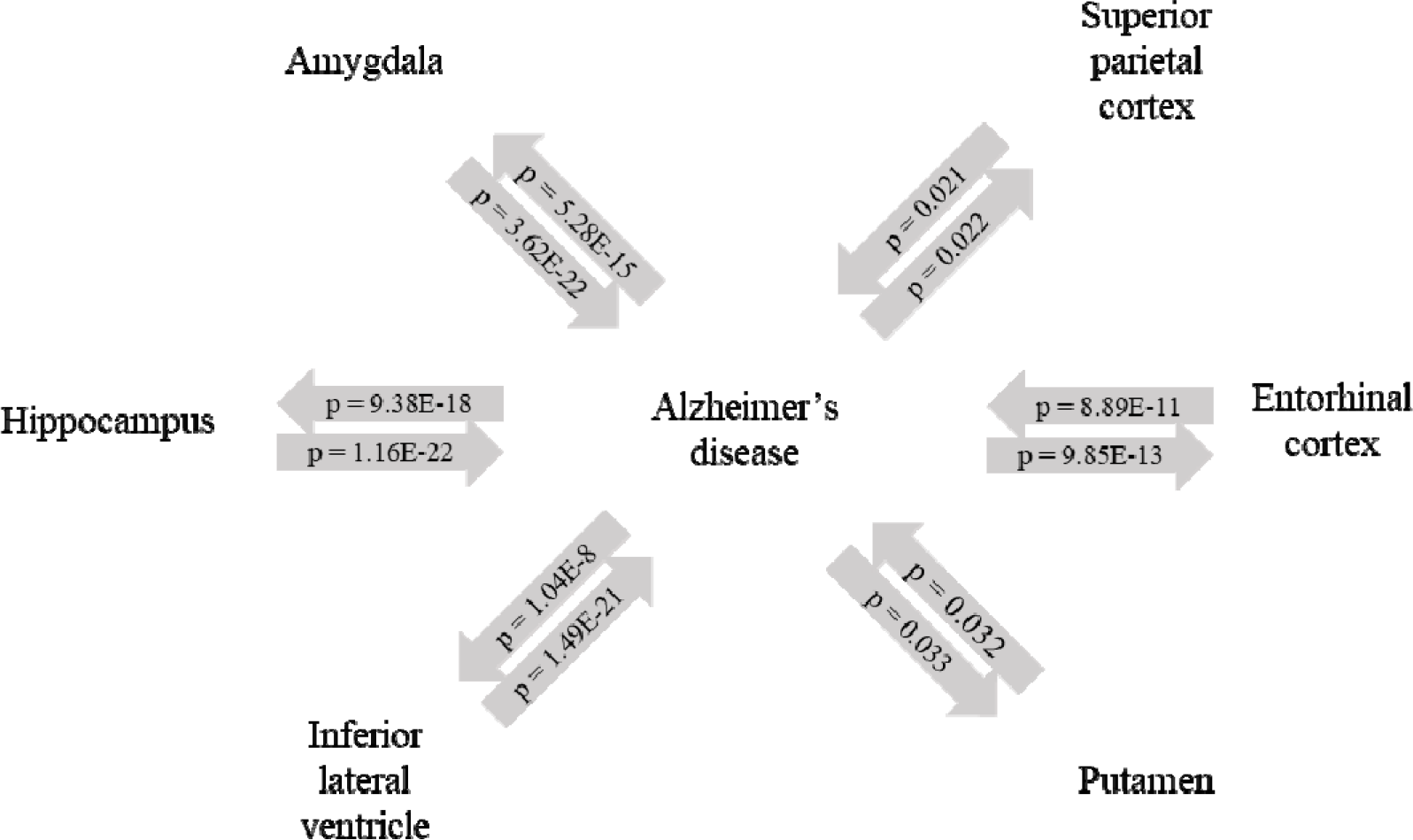
Results Mendelian randomization. In addition, LOAD significantly increased the superior parietal cortex but not *vice versa*. A significant Cochran’s Q statistic (p <0.001) for this model indicates a possible violation of the instrumental variable assumption. Leave-one-out analysis showed that excluding either *APOE*ε4 or rs75627662 from the analysis resulted in a non-significant association. In addition, exclusion of rs7384878 lead to a reduction in the width of the confidence interval.

### 4. Discussion

In this study, we aimed to determine whether effects of genetic variation on LOAD can be explained by regional brain morphology. To this aim, mediation analysis was performed using data collected by UKB and ADNI. We show a significant mediation effect of the entorhinal cortex, amygdala, hippocampus, inferior lateral ventricle and choroid plexus on the relation between *APOE* ε4 and LOAD. In addition, the effects of rs75627662 and rs7384878 on LOAD were mediated by the hippocampus and superior parietal cortex, respectively. MR was subsequently used to determine the directionality of the relationship between LOAD and the mediating brain regions. These results indicated a bidirectional relationship between the entorhinal cortex, hippocampus, amygdala, lateral inferior ventricle and choroid plexus, and LOAD.

We showed that *APOE* ε4 increased the risk of LOAD by altering the morphology of brain regions reported to be affected at an early stage in the pathological process of LOAD (12, 13, 50, 51). Our results correspond with previous research on the association of *APOE* ε4 with the middle temporal lobe, more specifically the entorhinal cortex, amygdala and hippocampus (26, 52). *APOE* ε4 has also been linked to enlargement of the choroid plexus, a hallmark of LOAD (53). Mediation analysis has been applied previously to study the determinants of LOAD. These studies differed from the current study by the variables used. Clinical scores or measures for cognitive function and/or behaviour were used as outcome measures and pathological hallmarks such as amyloid β and tau were used as mediator and independent variables (3, 54-61). Similar to our study, inclusion of brain measures as mediators identified the entorhinal cortex, hippocampus, amygdala and ventricles as mediators (55, 56, 60, 61). A mediation effect of the choroid plexus size on LOAD has not been reported previously.

Mendelian randomization pointed to a bidirectional effect between LOAD and the entorhinal cortex, hippocampus, amygdala, choroid plexus and the inferior lateral ventricle. From our results we can derive the following pathway regarding the effect of *APOE* ε4 on LOAD: The presence of *APOE* ε4 leads to a smaller entorhinal cortex, hippocampus and amygdala, and larger volume of the choroid plexus and inferior lateral ventricles. These differences in brain morphology increase the risk of developing LOAD, indicating that the effect of *APOE* ε4 is at the start of the disease pathology. The bidirectionality we found indicates that LOAD may lead to aggravation of the degeneration of the entorhinal cortex, hippocampus and amygdala, and expansion of the inferior lateral ventricle and choroid plexus. Multiple hypotheses have been raised regarding the pathological pathway by which *APOE* ε4 increases the risk of LOAD (57). ApoE plays a fundamental role in brain functioning by facilitating neuronal cholesterol metabolism, which is essential for axonal growth and synapse formation (33, 62). The apoE4 isoform was previously shown to lead to less efficient cholesterol transport and amyloid-β clearance, compared to the apoE2 and apoE3 isoforms (63). Accumulation of amyloid-β is one of the hallmarks of LOAD and plays a central role in the post-mortem diagnosis of LOAD and in many running theories regarding LOAD pathology (7). The choroid plexus has an important role in the production and removal of amyloid-β, and an enlargement of the choroid plexus has been associated with LOAD and was shown to be modified by *APOE* ε4 status (53). Possible explanations for the enlargement of the choroid plexus in LOAD were accumulation of amyloid-β and neuroinflammation, though it is not clear whether a larger choroid plexus is cause or consequence (53). Enlargement of the choroid plexus is also related to larger inferior ventricles.

In addition to *APOE* ε4, rs75627662 and rs7384878 were significantly related to LOAD. The effect of rs75627662was mediated by the entorhinal cortex, amygdala and hippocampus, and the effect of rs7384878 was mediated by the superior parietal cortex and putamen. Jansen et al. mapped rs75627662 to *APOE* and rs7384878 to *ZCWPW1* (17). The associations we found between rs75627662, the entorhinal cortex, amygdala and hippocampus, and LOAD might reflect the relations we found for *APOE* ε4. *ZCWPW1* encodes for a histone modification reader, involved in epigenetic regulation (64). In addition, *ZCWPW1* is in high linkage disequilibrium (LD) with multiple other genes including *PILRB, ZNF3, RFX3* and *NYAP1*. The effect of *ZCWPW1* on LOAD is likely to be an indirect effect via epigenetic modification of target genes or a reflection of a gene in LD (65). *PILRB* was shown to be expressed in multiple brain regions and expression levels were lower in LOAD cases compared to controls (66). *PILRB* is involved in regulation of the immune response (66). *ZNF3* is involved in a pathway resulting in protein degradation, oxidative stress, cell cycle control and tau pathology (67). Another hypothesis is that *ZCWPW1* activates binding of RFX3, thereby suppressing insulin resistance and decreasing the risk of LOAD (67). Lastly, *NYAP1* was shown in mice to affect brain size, neurite elongation and neuronal morphology (64). Mendelian randomization showed that the relationship between LOAD and the superior parietal cortex was unidirectional, with a higher risk of LOAD leading to an increased thickness of the superior parietal cortex. However, the Mendelian randomization model of the superior parietal cortex might be violating the instrumental variable assumption, making the validity of these results questionable. Based on our results we hypothesize that *APOE* ε4 induces LOAD via lipid metabolism, amyloid-β accumulation and inflammation. Secondary processes including protein degradation, oxidative stress and tau pathology, mediated via *ZCWPW1*, could lead to the spreading of neurodegeneration to other brain regions.

Several study limitations must be acknowledged. We found multiple, biologically plausible pathways of mediation in ADNI that were not replicated in UKB. This discrepancy between the two cohorts has multiple possible explanations. Due to a lack of UKB participants with a diagnosis of LOAD and MRI brain measures, a LOAD-by-proxy score was used instead of the LOAD status based on clinical diagnosis that was used in ADNI. The validity of the LOAD-by-proxy score as a substitute for clinical LOAD might be questioned. The LOAD-by-proxy score was used previously in GWAS, which showed a high genetic correlation between standard case-control status and the UKB by-proxy phenotype (17, 18, 27). However, the current study only used a subset of UKB participants and the proxy score may not be an appropriate substitute for clinical LOAD status (17, 68). Despite the high genetic correlation, true LOAD cases in the full UKB sample have higher polygenic risk scores compared to proxy cases (27) and inconsistent results have been attributed to the use of the LOAD proxy score (69). The LOAD-by-proxy score is sensitive to recall bias, because the score is based on self-reported parental information by the participants. Additionally, parental LOAD status may neither strongly correspond to the clinical LOAD status of the participant (19, 68, 70, 71) nor indicate underlying pathophysiology. A second explanation for the differing results from ADNI and UKB is that these are different types of cohorts. ADNI is a case-control cohort in which LOAD cases were compared to controls that were generally healthy, predominantly of European White ancestry and overwhelmingly highly educated participants (29, 72), with likely greater cognitive reserve (and thereby possibly delayed LOAD onset), thereby limiting the generalizability of the results. Substantial differences between ADNI and community based cohorts were reported previously (72). UKB participants have been shown to be overly healthy compared to the general population (73), raising the possibility of a higher resilience to the influence of genetic risk factors associated with LOAD. Additionally, UKB participants were on average slightly younger than the ADNI participants. Only participants that responded to the invitation for a second assessment underwent MRI, enhancing the possibility of healthy volunteer bias. Lastly, MRI measures from UKB were obtained using FreeSurfer version 5.3 whereas MRI scans from ADNI were processed by version 5.1. Different versions of FreeSurfer have been compared by others, who concluded that there were significant differences in the absolute values of the measures obtained (74, 75). However, results from correlation studies as well as the ability to differentiate between cases and controls were comparable. It is therefore possible to compare the results obtained from UKB and ADNI (74, 75).

Our findings demonstrate that combining genetic and neuroimaging data can give insight into the causal neuropathological pathways of LOAD. Future studies may expand the number of genetic risk variants under investigation or combine the effects of multiple SNPs into a polygenic risk score. Through this study we illustrated that mediation analysis combined with MR, enabling statements about causality, may increase knowledge on the (order of) pathological processes underlying LOAD. This can be valuable for developing tools for optimized diagnosis and treatment.

## Data Availability

This study made use of UKB data, obtained from the repository under accession code 55392, and from ADNI data, obtained via the account of the corresponding author.

## Acknowledgement

This study made use of UKB data, obtained from the repository under accession code 55392.

Data collection and sharing for this project was funded by the Alzheimer’s Disease Neuroimaging Initiative (ADNI) (National Institutes of Health Grant U01 AG024904) and DOD ADNI (Department of Defense award number W81XWH-12-2-0012). ADNI is funded by the National Institute on Aging, the National Institute of Biomedical Imaging and Bioengineering, and through generous contributions from the following: AbbVie, Alzheimer’s Association; Alzheimer’s Drug Discovery Foundation; Araclon Biotech; BioClinica, Inc.; Biogen; Bristol-Myers Squibb Company; CereSpir, Inc.; Cogstate; Eisai Inc.; Elan Pharmaceuticals, Inc.; Eli Lilly and Company; EuroImmun; F. Hoffmann-La Roche Ltd and its affiliated company Genentech, Inc.; Fujirebio; GE Healthcare; IXICO Ltd.; Janssen Alzheimer Immunotherapy Research & Development, LLC.; Johnson & Johnson Pharmaceutical Research & Development LLC.; Lumosity; Lundbeck; Merck & Co., Inc.; Meso Scale Diagnostics, LLC.; NeuroRx Research; Neurotrack Technologies; Novartis Pharmaceuticals Corporation; Pfizer Inc.; Piramal Imaging; Servier; Takeda Pharmaceutical Company; and Transition Therapeutics. The Canadian Institutes of Health Research is providing funds to support ADNI clinical sites in Canada. Private sector contributions are facilitated by the Foundation for the National Institutes of Health (www.fnih.org). The grantee organization is the Northern California Institute for Research and Education, and the study is coordinated by the Alzheimer’s Therapeutic Research Institute at the University of Southern California. ADNI data are disseminated by the Laboratory for Neuro Imaging at the University of Southern California.

