## Supplementary info for "Brain morphology mediating the effect of genetic risk variants on Alzheimer’s disease"

Supplement

### Supplementary table 1a: Variable description UK Biobank.

| Variable | Description | Fieldcode UK Biobank |
| --- | --- | --- |
| **AD proxy score** |  |  |
| Participant AD diagnosis | Based on presence of ICD-10 codes G30 or F00, or report of AD. | 41270, 42020 |
| Paternal AD diagnosis | Self-reported by participants. | 20107 |
| Paternal age (at death) | Reported by participant. | 29,461,807 |
| Maternal AD diagnosis | Self-reported by participants. | 20110 |
| Maternal age (at death) | Reported by participant. | 18,453,526 |
| **Demographics** |  |  |
| Age | Age filled in at the time of assessment 2 (visit when imaging data was collected) | 21003 |
| Sex | Based on the sex based on genotyping. |  |
| Ethnicity | Self-identified as 'White British' and very similar genetic ancestry based on a principal components analysis of the genotypes. | 22006 |
| Site | UK Biobank assessment centre | 54 |
| **Brain measures** |  |  |
| Cortical area | Freesurfer DKT | 27143-27173 |
|  |  | 27236-27266 |
|  | Freesurfer desikan white | 26721, 26822, |
|  |  | 26752, 26853, |
|  |  | 26722, 26823 |
|  | Freesurfer a2009s | 27371, 27593 |
| Cortical thickness | Freesurfer DKT | 27174-27204 |
|  |  | 27267-27297 |
|  | Freesurfer desikan white | 26755, 26856, |
|  |  | 26786, 26887, |
|  |  | 26756, 26857 |
|  | Freesurfer a2009s | 27445, 27667 |
| Subcortical volume | Freesurfer ASEG | 26514-26537 |
|  |  | 26552-26567 |
|  |  | 26583-26598 |

### Supplementary table 1b: Variable description ADNI.

| Variable | Description | Table ADNI | Fieldcode ADNI |
| --- | --- | --- | --- |
| **AD** |  |  |  |
| Diagnosis ADNI1 | 1=NL; 2=MCI; 3=AD | DXSUM | DXCURREN |
| Diagnosis ADNIGO/2 | 1=Stable: NL to NL; 2=Stable: MCI to MCI; 3=Stable: Dementia to Dementia; 4=Conversion: NL to MCI; 5=Conversion: MCI to Dementia; 6=Conversion: NL to Dementia; 7=Reversion: MCI to NL; 8=Reversion: Dementia to MCI; 9=Reversion: Dementia to NL |  | DXCHANGE |
| **Demographics** |  |  |  |
| Year of birth | Participant year of birth | PTDEMOG | PTDOBYY |
| Date of imaging | Examination Date | UCSFFSX51 | EXAMDATE |
| Sex/gender | 1=Male; 2=Female | PTDEMOG | PTGENDER |
| Race | 1=American Indian or Alaskan Native; 2=Asian; 3=Native Hawaiian or Other Pacific Islander; 4=Black or African American; **5=White**; 6=More than one race; 7=Unknown | PTDEMOG | PTRACCAT |
| Site |  |  | SITEID |
| **Brain measures** |  |  |  |
| Cortical area | ADNI1/GO/2: 3T  (Freesurfer 5.1) | UCSFFSX51 | ST13SA-ST15SA, ST23SA-ST26SA, ST31SA, ST32SA, ST34SA-ST36SA, ST38SA-ST40SA, ST43SA-ST52SA, ST54SA-ST60SA, ST62SA, ST72SA-ST74SA, ST82SA-ST85SA, ST90SA, ST91SA, ST93SA-ST95SA, ST97SA-ST99SA, ST102SA-ST111SA, ST113SA-ST119SA, ST121SA, ST129SA, ST130SA |
| Cortical thickness | ADNI1/GO/2: 3T  (Freesurfer 5.1) | UCSFFSX51 | ST13TA-ST15TA, ST23TA-ST26TA, ST31TA, ST32TA, ST34TA-ST36TA, ST38TA-ST40TA, ST43TA-ST52TA, ST54TA-ST60TA, ST62TA, ST72TA-ST74TA, ST82TA-ST85TA, ST90TA, ST91TA, ST93TA-ST95TA, ST97TA-ST99TA, ST102TA-ST111TA, ST113TA-ST119TA, ST121TA, ST129TA, ST130TA |
| Subcortical volume | ADNI1/GO/2: 3T,  (Freesurfer 5.1) | UCSFFSX51 | ST1SV-ST9SV, ST11SV, ST12SV, ST16SV-ST18SV,ST21SV,ST29SV,ST30SV, ST37SV, ST42SV, ST53SV, ST61SV, ST65SV, ST68SV-ST71SV, ST75SV-ST77SV, ST80SV, ST88SV, ST89SV, ST96SV, ST101SV, ST112SV, ST120SV, ST124SV, ST125SV, ST127SV, ST128SV, ST147SV, ST148SV, ST150SV, ST151SV, ST153SV-ST155SV |

Supplementary table 2: SNP selection.
Lead SNPs for each genomic risk loci obtained by FUMA. Not all participants had information on all genetic variants, participants were excluded in a model based manner based on the variant included.

| GenomicLocus | uniqID | rsID | chr | pos | start | end | UKB sample size |
| --- | --- | --- | --- | --- | --- | --- | --- |
| 1 | 1:207750568:C:T | rs679515 | 1 | 207750568 | 207679307 | 207806730 | 38,845 |
| 2 | 2:127891427:A:C | rs4663105 | 2 | 127891427 | 127826533 | 127894851 | 37,728 |
| 3 | 3:57226150:C:T | rs184384746 | 3 | 57226150 | 56252241 | 57879720 | na |
| 4 | 4:11024682:C:G | rs6448451 | 4 | 11024682 | 11014822 | 11041549 | 38,897 |
| 5 | 6:40942196:A:G | rs187370608 | 6 | 40942196 | 40706366 | 41129252 | na |
| 6 | 7:99932049:C:T | rs7384878 | 7 | 99932049 | 99777422 | 100091795 | 38,325 |
| 7 | 7:145950029:C:T | rs114360492 | 7 | 145950029 | 145181183 | 146573693 | na |
| 8 | 8:27466315:C:T | rs1532278 | 8 | 27466315 | 27456253 | 27468503 | 38,922 |
| 9 | 11:60021948:A:G | rs1582763 | 11 | 60021948 | 59826677 | 60099912 | 38,853 |
| 10 | 11:85850243:C:T | rs3844143 | 11 | 85850243 | 85652251 | 85869737 | 38,922 |
| 11 | 17:4984447:A:G | rs9916042 | 17 | 4984447 | 4958842 | 5013491 | 38,508 |
| 12 | 19:1053524:C:G | rs3752241 | 19 | 1053524 | 1053524 | 1053524 | 38,176 |
| 13 | 19:45413576:C:T | rs75627662 | 19 | 45413576 | 44724661 | 46491516 | 38,771 |

na: no participant had information available for this snp

Supplementary table 3: Total effect genetic variants on LOAD.
Total effect of the genetic variants on the AD proxy score (UK Biobank) and the odds of AD (ADNI), corrected for age and sex.

|  | UK Biobank | | ADNI | | |
| --- | --- | --- | --- | --- | --- |
| Genetic variant | Estimate | p-value | Estimate | OR | p-value |
| apoe4 | 0.105 | 8.97E-146 | 1.339 | 3.817 | 9.85E-15 |
| rs75627662 | 0.050 | 4.96E-43 | 0.736 | 2.088 | 2.48E-06 |
| rs4663105 | 0.011 | 2.36E-04 | 0.159 | 1.172 | 0.221 |
| rs7384878 | -0.011 | 0.001 | -0.392 | 0.676 | 0.009 |
| rs679515 | 0.012 | 0.002 | 0.087 | 1.091 | 0.605 |
| rs1582763 | -0.009 | 0.002 | -0.144 | 0.866 | 0.275 |
| rs1532278 | -0.009 | 0.003 | -0.246 | 0.782 | 0.083 |
| rs3844143 | -0.009 | 0.003 | -0.057 | 0.944 | 0.671 |
| rs6448451 | 0.006 | 0.082 | 0.139 | 1.149 | 0.341 |
| rs9916042 | 0.003 | 0.317 | 0.291 | 1.337 | 0.040 |
| rs3752241 | -0.002 | 0.626 | -0.110 | 0.896 | 0.532 |

### Supplementary table 5a: Results mendelian randomization analysis.

| Brain measure | Direction | IVW | | | MR-Egger | | | Weighted median | | |
| --- | --- | --- | --- | --- | --- | --- | --- | --- | --- | --- |
|  |  | Beta | SE | P-value | Beta | SE | P-value | Beta | SE | P-value |
| Entorhinal thickness | → | -3.456 | 0.533 | **8.89E-11** | -3.994 | 0.598 | **2.32E-11** | -3.542 | 0.764 | **3.51E-06** |
|  | ← | -0.239 | 0.033 | **9.85E-13** | -0.267 | 0.046 | **4.65E-09** | -0.256 | 0.050 | **3.04E-07** |
| Amygdala | → | -3.088 | 0.319 | **3.62E-22** | -3.369 | 0.439 | **1.69E-14** | -3.219 | 0.614 | **1.59E-07** |
|  | ← | -0.303 | 0.039 | **5.28E-15** | -0.303 | 0.053 | **8.60E-09** | -0.307 | 0.058 | **1.12E-07** |
| Hippocampus | → | -2.870 | 0.293 | **1.16E-22** | -2.774 | 0.394 | **1.82E-12** | -3.001 | 0.547 | **4.02E-08** |
|  | ← | -0.330 | 0.038 | **9.38E-18** | -0.327 | 0.052 | **4.33E-10** | -0.323 | 0.060 | **5.95E-08** |
| Inferior Lateral Ventricle | → | 3.993 | 0.419 | **1.49E-21** | 3.942 | 0.590 | **2.40E-11** | 4.219 | 1.051 | **0.0001** |
|  | ← | 0.228 | 0.040 | **1.04E-08** | 0.224 | 0.054 | **3.51E-05** | 0.232 | 0.054 | **1.71E-05** |
| Putamen | → | -2.999 | 1.402 | **0.032** | -0.242 | 0.137 | 0.078 | -1.706 | 3.747 | 0.649 |
|  | ← | -0.096 | 0.045 | **0.033** | -0.041 | 0.029 | 0.156 | -0.058 | 0.046 | 0.213 |
| Superior parietal (ukb) | → | 1.746 | 0.759 | **0.021** | 2.184 | 1.298 | 0.092 | 0.852 | 0.556 | 0.126 |
|  | ← | 0.198 | 0.087 | **0.022** | 0.240 | 0.108 | **0.027** | 0.221 | 0.046 | **1.28E-06** |

### Supplementary table 5b: Results diagnostics measures mendelian randomization analysis.

| Brain measure | Direction | MR-Egger (intercept) | | | I²-statistic | Cochran's Q test | |
| --- | --- | --- | --- | --- | --- | --- | --- |
|  |  | Beta | SE | P-value |  | Estimate | p-value |
| Entorhinal thickness | → | 0.112 | 0.069 | 0.108 | 0.728 | 15.021 | 0.090 |
|  | ← | 0.021 | 0.023 | 0.361 | 0.818 | 8.311 | 0.503 |
| Amygdala | → | 0.057 | 0.061 | 0.352 | 0.667 | 6.060 | 0.734 |
|  | ← | 0.000 | 0.027 | 0.987 | 0.819 | 3.328 | 0.950 |
| Hippocampus | → | -0.022 | 0.060 | 0.717 | 0.729 | 4.538 | 0.873 |
|  | ← | -0.002 | 0.027 | 0.931 | 0.819 | 2.771 | 0.973 |
| Inferior Lateral Ventricle | → | 0.008 | 0.063 | 0.898 | 0.438 | 9.714 | 0.374 |
|  | ← | 0.003 | 0.027 | 0.918 | 0.817 | 2.707 | 0.975 |
| Putamen | → | -0.242 | 0.137 | 0.078 | 0.080 | 51.321 | **6.07E-08** |
|  | ← | -0.041 | 0.029 | 0.156 | 0.815 | 10.650 | 0.300 |
| Superior parietal (ukb) | → | -0.006 | 0.013 | 0.670 | 0.562 | 591.96 | **1.11E-121** |
|  | ← | -0.002 | 0.003 | 0.497 | 0.983 | 39.381 | **9.83E-06** |

N = 505,502 UKB participants

N = 1740 ADNI 1/GO/2 participants

n = 39,565 White British race

5,984 excluded

n = 780 White British race

9 excluded

n = 38,922 with parental AD status and age (at death)

643 excluded

n = 476 AD or cognitively normal

304 excluded

n = 46,852 with MRI data

465,447 excluded

n = 1068 with MRI data

672 excluded

n = 45,549 were genotyped and had information on at least one SNPs of interest

1,303 excluded

n = 789 were genotyped and had information on the SNPs of interest

279 excluded

### Supplementary figure 1: Overview inclusion and exclusion of UK Biobank and ADNI participants.

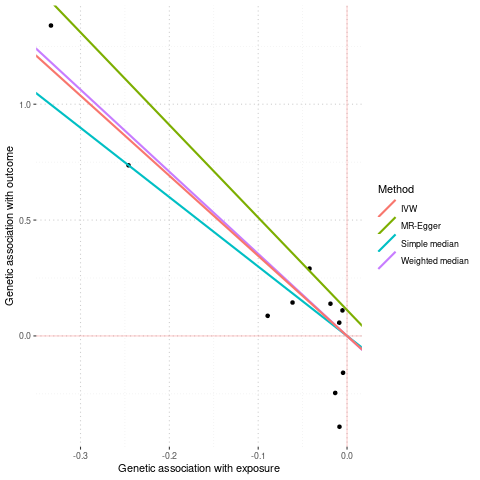

A

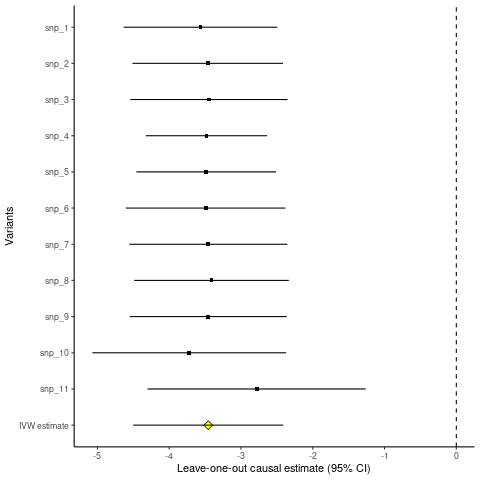

B

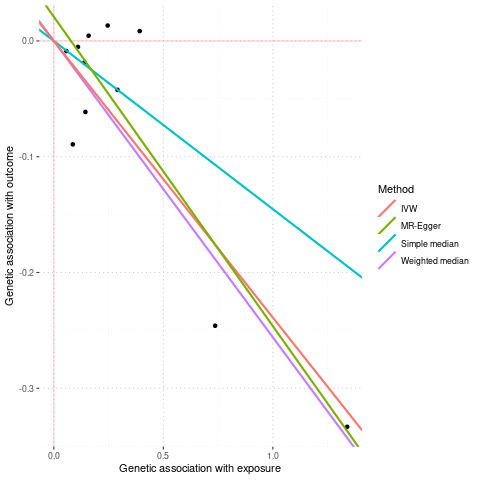

C

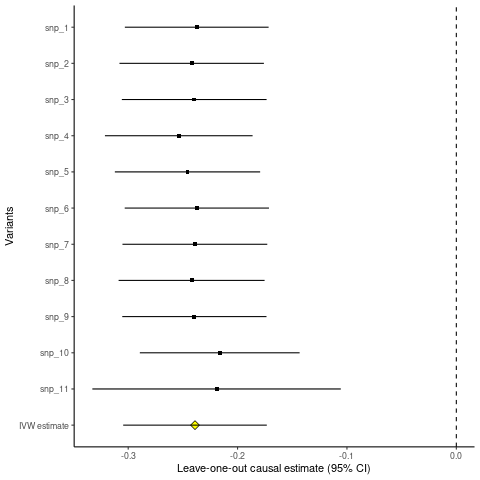

D

Supplementary figure 2a: Diagnostics plots MR entorhinal cortex.
Plots A and B show the diagnostics of the MR analysis and plot C and D from the reverse MR. Plot A and C show the genetic association with the outcome and the exposure calculated using the different methods. Plot B and D show the estimates and 95% CI from the leave-one-out analysis compared to the overall IVW estimate.

A

B

C

D

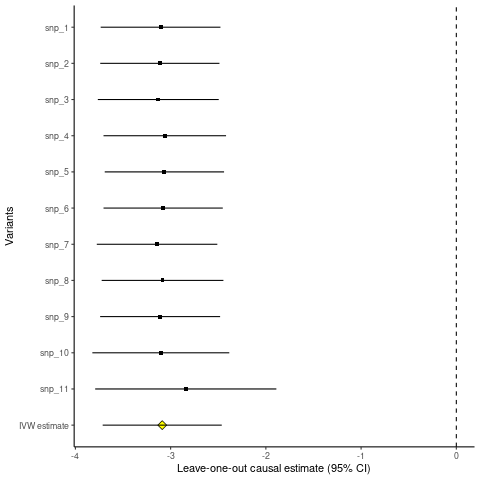

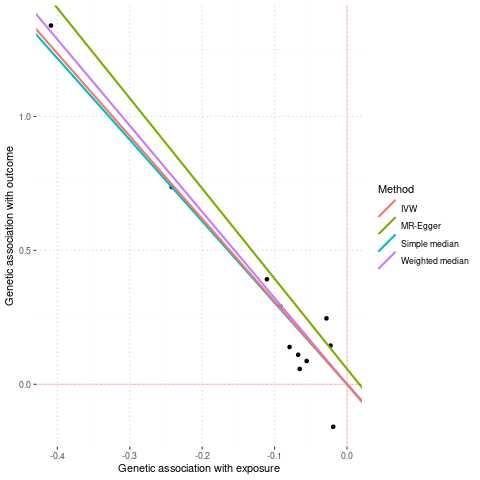

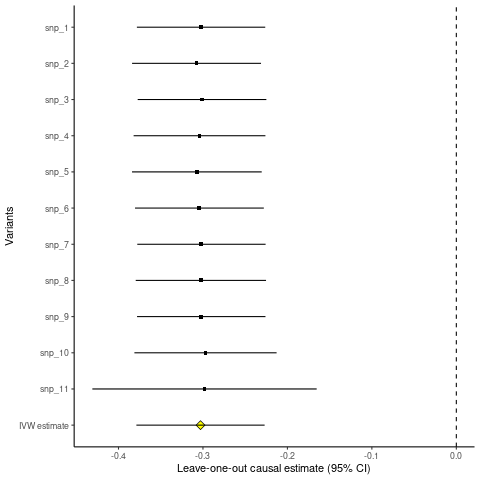

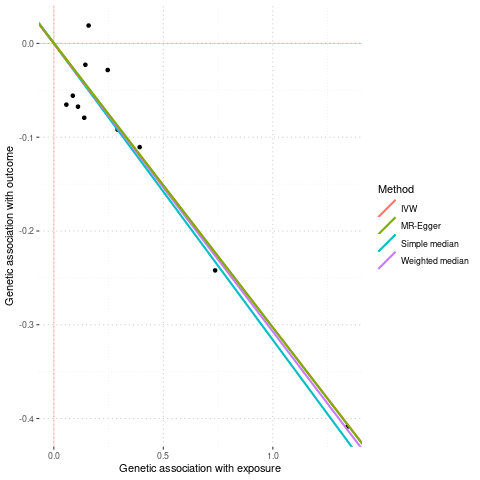

Supplementary figure 2b: Diagnostics plots MR amygdala.
Plots A and B show the diagnostics of the MR analysis and plot C and D from the reverse MR. Plot A and C show the genetic association with the outcome and the exposure calculated using the different methods. Plot B and D show the estimates and 95% CI from the leave-one-out analysis compared to the overall IVW estimate.

A

B

C

D

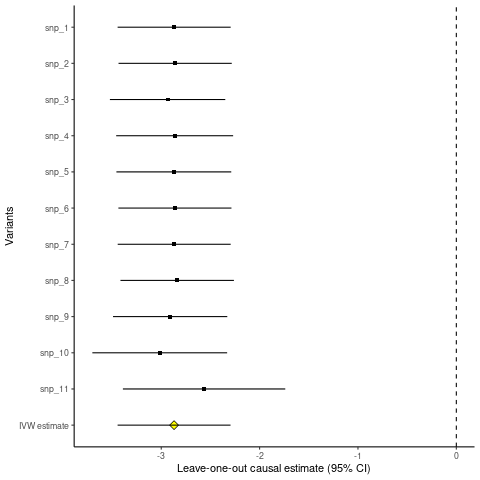

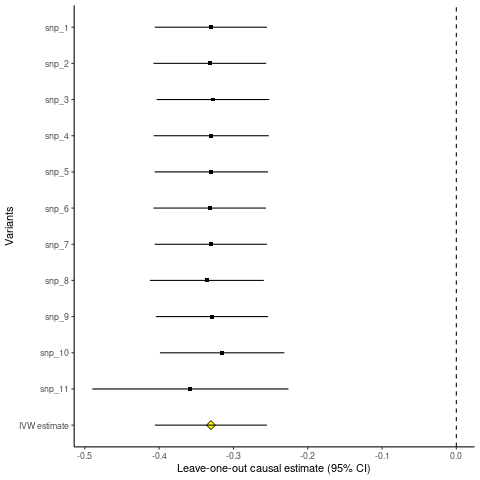

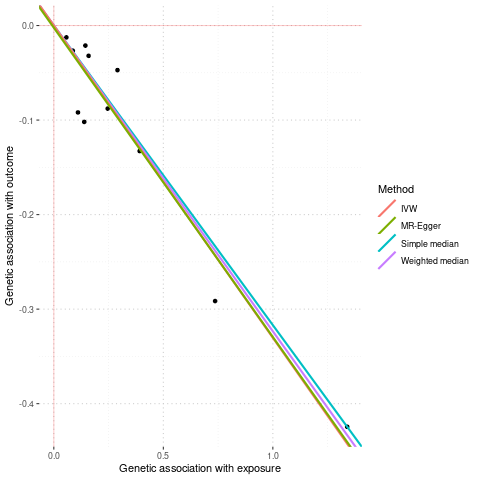

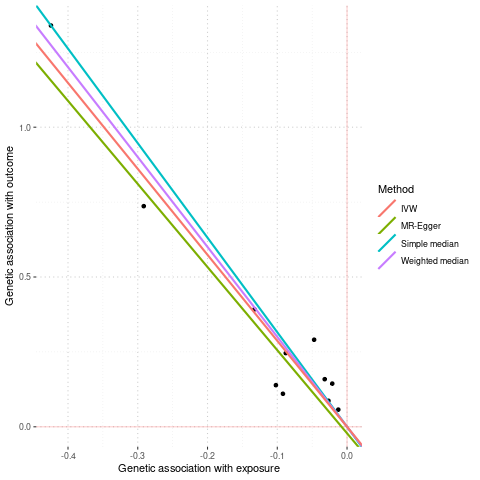

Supplementary figure 2c: Diagnostics plots MR hippocampus.
Plots A and B show the diagnostics of the MR analysis and plot C and D from the reverse MR. Plot A and C show the genetic association with the outcome and the exposure calculated using the different methods. Plot B and D show the estimates and 95% CI from the leave-one-out analysis compared to the overall IVW estimate.

A

B

C

D

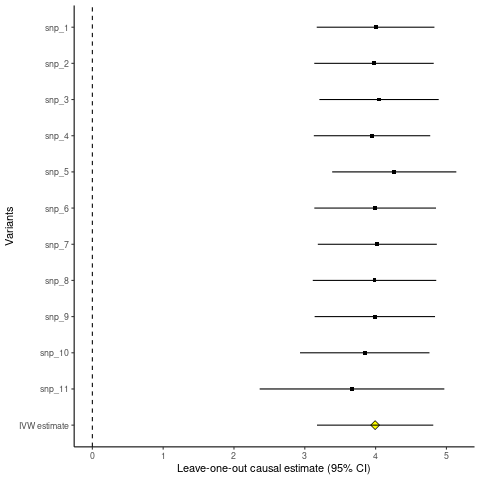

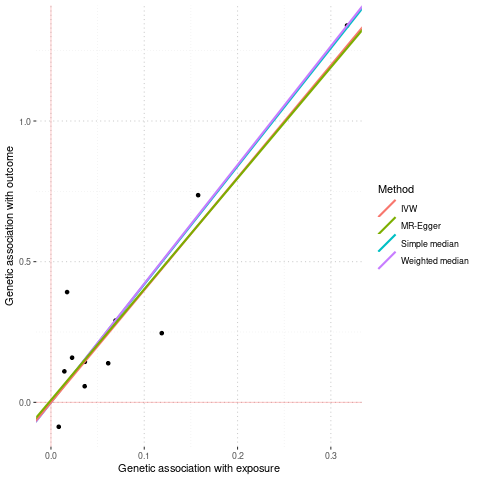

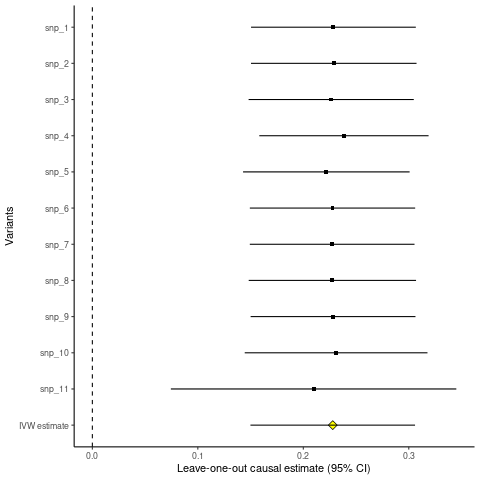

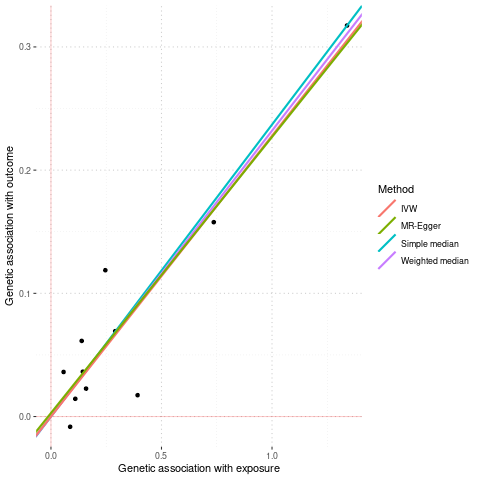

Supplementary figure 2d: Diagnostics plots MR inferior lateral ventricle.
Plots A and B show the diagnostics of the MR analysis and plot C and D from the reverse MR. Plot A and C show the genetic association with the outcome and the exposure calculated using the different methods. Plot B and D show the estimates and 95% CI from the leave-one-out analysis compared to the overall IVW estimate.

A

B

C

D

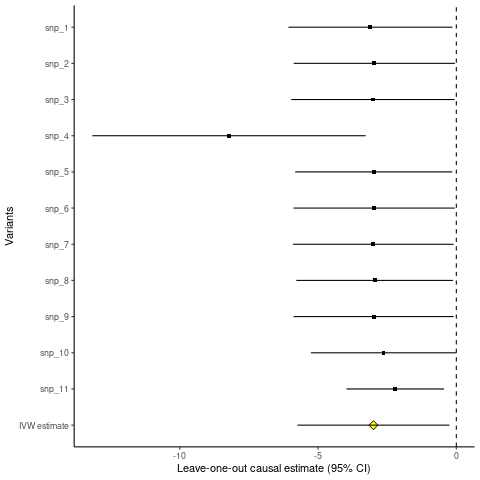

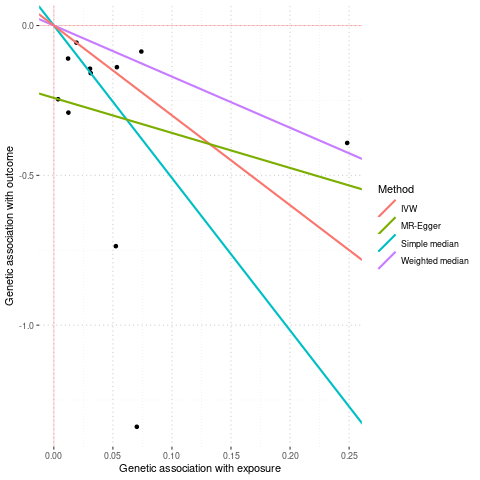

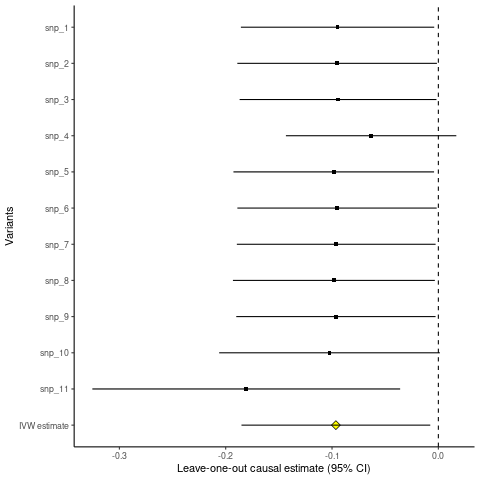

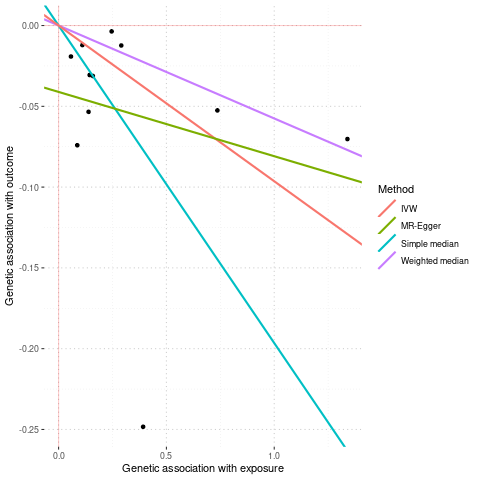

Supplementary figure 2e: Diagnostics plots MR putamen.
Plots A and B show the diagnostics of the MR analysis and plot C and D from the reverse MR. Plot A and C show the genetic association with the outcome and the exposure calculated using the different methods. Plot B and D show the estimates and 95% CI from the leave-one-out analysis compared to the overall IVW estimate.

A

B

C

D

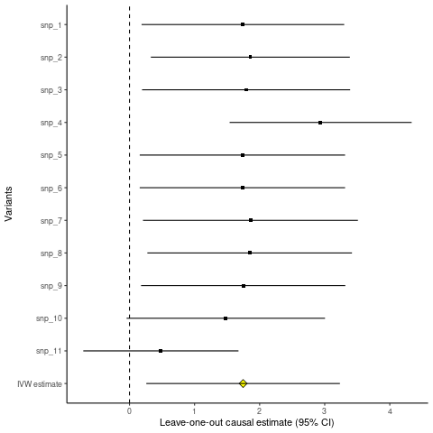

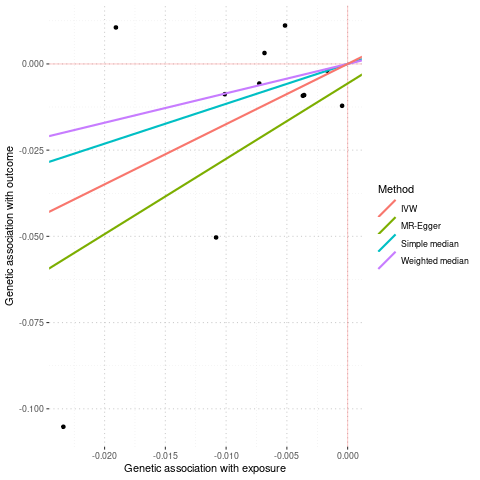

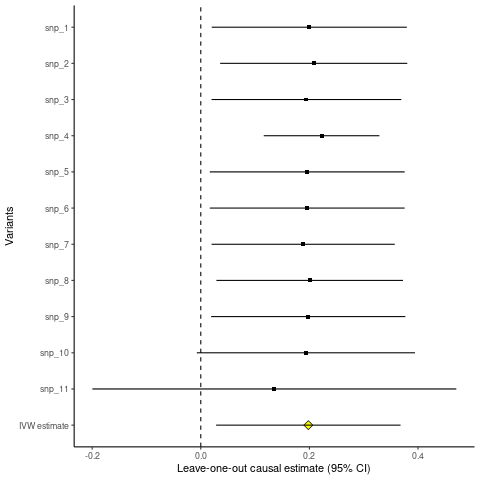

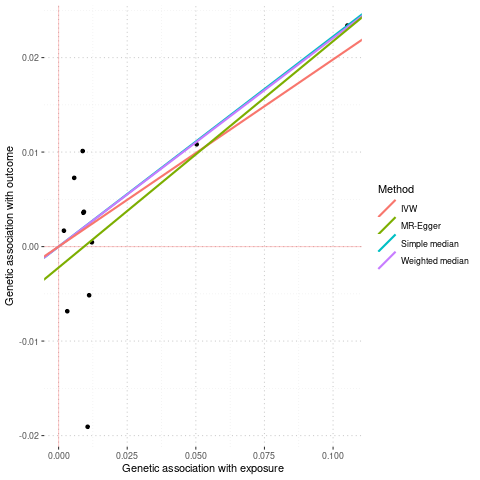

Supplementary figure 2f: Diagnostics plots MR superior parietal cortex.
Plots A and B show the diagnostics of the MR analysis and plot C and D from the reverse MR. Plot A and C show the genetic association with the outcome and the exposure calculated using the different methods. Plot B and D show the estimates and 95% CI from the leave-one-out analysis compared to the overall IVW estimate.
